# A snowball approach for screening of asymptomatic Rheumatic Heart Disease among school age children in Ethiopia: A comparative cross-sectional study

**DOI:** 10.1101/2025.09.24.25336540

**Authors:** Demeke Mekonnen, Tigist Mekonnen, Elisabeth Lilian Pia Sattler, Mahlet Worku Molla, Bamba Gaye, Tigist Workneh Leulseged

## Abstract

**Background:** Rheumatic heart disease (RHD) is a common complication of acute rheumatic fever (ARF). ARF and RHD are the leading causes of cardiovascular morbidity and mortality. Early detection and structured prevention programs can halt the devastating effects of RHD. The study aimed to determine the overall prevalence of asymptomatic RHD and asses the case detection rates between random screening and snow ball approaches at selected schools in Addis Ababa, Ethiopia.

**Methods:** A community based, comparative, cross-sectional study was conducted at five elementary schools in Addis Ababa. Two groups of study population were identified as positive contact-known RHD patient in their class and negative contact-no known contact in their class. RHD screening was done using World heart federation 2012 echocardiographic criteria (as borderline and definitive RHD) by Lumify handheld device. Data was summarized using frequencies with percentages and median with interquartile range. Multivariable binary logistic regression model was run and adjusted odds ratio (AOR) was used to measure the strength.

**Results:** A total of 265 children (positive-108 and negative-157 contact) were screened. The median age of the participants was 11 years. The overall prevalence of asymptomatic RHD was 4.9%. The stratified prevalence among children with negative contact history-3/157 (1.9%) and positive contacts was 10/108 (9.3%). Out of the 10 cases, 2 students had mild mitral regurgitation. After adjusting for age and sex, the odds of developing asymptomatic RHD among participants with positive contact history was 5.67 times than participants with negative contact history (AOR= 5.67, 95% CI= 1.40, 22.98, p-value=0.015).

**Conclusion:** The prevalence of asymptomatic RHD was higher among children with positive contact history. The majority of these children were diagnosed with definitive RHD as compared to those with negative contact history. Screening using a snowball technique gives us a superior yield which needs further study with larger sample.

**Key messages:** RHD is the leading cause of morbidity and mortality in the developing countries. Early detection of asymptomatic RHD can help prevent progression of the disease. Screening of school children helps detect RHD early though costly and unreachable. Integration of snowball approach to trace contacts of RHD patients will minimize cost and improves detection rate.

## Introduction

Acute rheumatic fever (ARF) is a delayed, non-suppurative sequelae of a pharyngeal infection with Group A β -hemolytic streptococcus (GAS). Following the initial pharyngitis, a latent period of two to three weeks occurs before the first signs or symptoms of ARF appear. The disease presents with various manifestations that may include arthritis, carditis, chorea, subcutaneous nodules, and erythema marginatum (1). Globally there are over 33 million people with RHD with a record of 470,000 new cases and nearly 320,000 deaths each year. Of these, most occur in developing countries affecting millions of people and leading to cardiovascular death during the first 5 decades of life due to the high transmission rate of the bacteria in overcrowded living situations including schools (2-4).

The initiation of early secondary prophylaxis significantly improves the long-term outcome of RHD and is an effective strategy for prevention (12-14). However, among the challenges associated with early diagnosis and timely intervention is that only two-thirds of patients with ARF will report a preceding sore throat, only 40%–60% of ARF cases progress to RHD, and up to 75% of children with RHD have no memory of symptoms consistent with previous ARF (2, 8). Therefore, most cases of RHD are not necessarily typified by the classic sequence of GAS pharyngitis resulting in symptomatic ARF with progression to RHD, suggesting that rheumatic carditis frequently occurs at a subclinical level (9).

According to school screening programs, it is reported that the prevalence of asymptomatic RHD in sub-Saharan Africa is 0.5 to 3% (5-7). A multicenter school screening conducted in 2016 for children revealed that the prevalence of definite RHD was 14/1000 and borderline RHD was 5/1000 (5). Considering the disease burden in the country, this is a low yield which might be attributable to the random screening method used. Hence, designing a systematic screening approach that can lead to detection of a large number of cases is essential for early intervention and better outcome.

Therefore, the objective of this study was to determine the prevalence of asymptomatic RHD using random and snowball screening methods and the association of screening methods with case detection rate among children at selected schools in Addis Ababa, Ethiopia.

## Materials and Methods

### Study Design, Population and Sample size

A community based comparative cross-sectional study, following the STROBE guidelines for analytical cross-sectional studies, was conducted at five elementary and phase I secondary schools, in Addis Ababa, Ethiopia, from January to June, 2022. The comparator groups were the following;

- Snowball approach: children who are close contacts of a known case of RHD (**Positive contact**): Three index cases (2 boys and 1 girl) were selected from a follow-up clinic and contacts were traced from their schools. In the three schools, classrooms of the index cases and close contacts were identified for screening. A total of 123 children were identified.
- Random screening: no known contact of known or symptomatic RHD patient (**Negative contact-Control group**): Two other schools were selected from the same woreda (district) for random screening of 200 students.

Children who were already diagnosed with heart disease or had clinical symptoms of heart disease were excluded (symptomatic RHD). After excluding the ineligible children, a total of 265 children (108 -positive contact history and 157 -negative contact) were included in the study. (Figure 1)

**Fig. 1.**
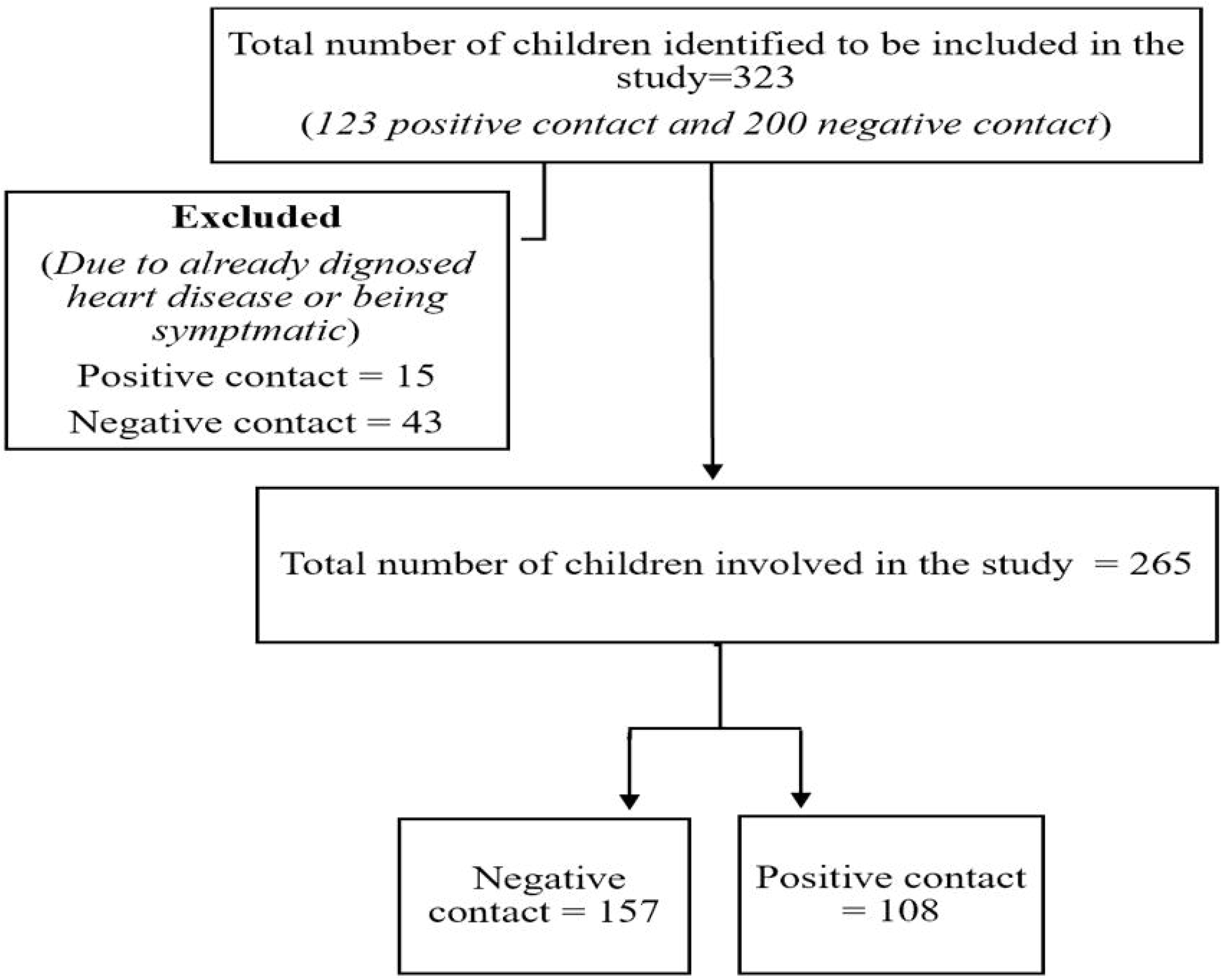

A post-hoc power analysis was calculated using G^*^Power 3.19.4 to check the power of the study using a two-tailed z-test for difference between two independent proportions with the following statistical parameters; 5% level of significance, prevalence and sample size in the positive contact group of 9.3% and 108, respectively; and prevalence and sample size in the negative contact group of 1.9% and 157, respectively. Finally, the power of the study was found to be 75.8%.

### Operational Definitions

#### Asymptomatic rheumatic heart disease

Children with RHD diagnosis based on the world heart federation echocardiographic criteria but do not have any symptoms (15).

#### Borderline RHD

- Echocardiographic features in an individual aged ≤20 years which are abnormal but do not fulfill criteria for the diagnosis of RHD according to WHF. Borderline RHD on echocardiogram without a documented history of ARF.

#### Definite RHD

- Echocardiographic features in an individual of any age which are abnormal and fulfill criteria for the diagnosis of RHD according to the WHF criteria. Echocardiographic changes that meet the criteria for ‘definite RHD’ are considered to be rheumatic in origin, provided that other etiologies have been excluded by echocardiography and clinical context.

### Data Collection Procedures and Quality Assurance

Ethical clearance was taken from St. Peter hospital research directorate and community discussion was conducted with the school heads and representatives. After consent was taken from the families, data was captured and analyzed after personal deidentification. Basic and clinical data was obtained from the eligible participants by trained BSc nurses and a General Practitioner. Screening for asymptomatic RHD as per the WHF criteria was made using a Lumify handheld echocardiography (10, 11) by an experienced consultant pediatric cardiologist. When there was a suspected case, confirmation was made using standard echo machine GE-vivid e-9. For the positive cases, management was initiated according to the guideline and they were enrolled for a subsequent follow up appointment at the pediatric cardiac unit at St. Peter specialized hospital every three months.

Data consistency and completeness were checked before an attempt was made to enter the code and analyze the data. Data cleaning through checking for inconsistencies, numerical errors and missing parameters was done. Once data cleaning was complete, data was exported to SPSS version 25.0 software for analysis.

### Statistical Analysis

Data for categorical variables was summarized using frequencies with percentages. Data for the numeric variable (age) was summarized using median (Interquartile Range) due to the skewed distribution of the data (Kolmogorov Smirnov and Shapiro-wilk tests of normality, both p-value <0.0001).

To check for the presence of a statistically significant differences in the characteristics of the two comparator groups (positive and negative contacts), a chi-square test and Fisher’s exact test were used.

Multivariate analysis was done for association and for significant values, adjusted odds ratio (AOR) with its 95% confidence interval (CI) was used to measure the strength of the relationship. The adequacy of the final model was assessed using the Hosmer and Leme show goodness of fit test and the final model fitted the data well (X^2^_(4)_ = 1.463 and p-value = 0.833).

## Results

### Baseline characteristics of the participants

The median age of the participants was 11 years (IQR, 9-14) and the majority (79.6%) were males. All of the participants claimed that they had an episode of acute tonsillopharyngitis (ATP) in the past three months and for which all of them were treated with antibiotics. Of which only 203 (76.6%) claimed that they have received the treatment within 2 weeks of initial symptoms, although there was no specific report of which antibiotic, what dose and for how long apart from at least 7 days of treatment. The participants were asked if they are aware that untreated ATP might result in heart disease and they all responded no.

Comparisons between the two study groups revealed a significant difference based on their age only. Accordingly, a significantly larger proportion (77.1%) of participants with negative contact were in the age group of 8-12 years while the majority (61.1%) of those with positive contact were in the age range of 13-15 years (p-value < 0.0001). (Table 1)

**Table 1:**
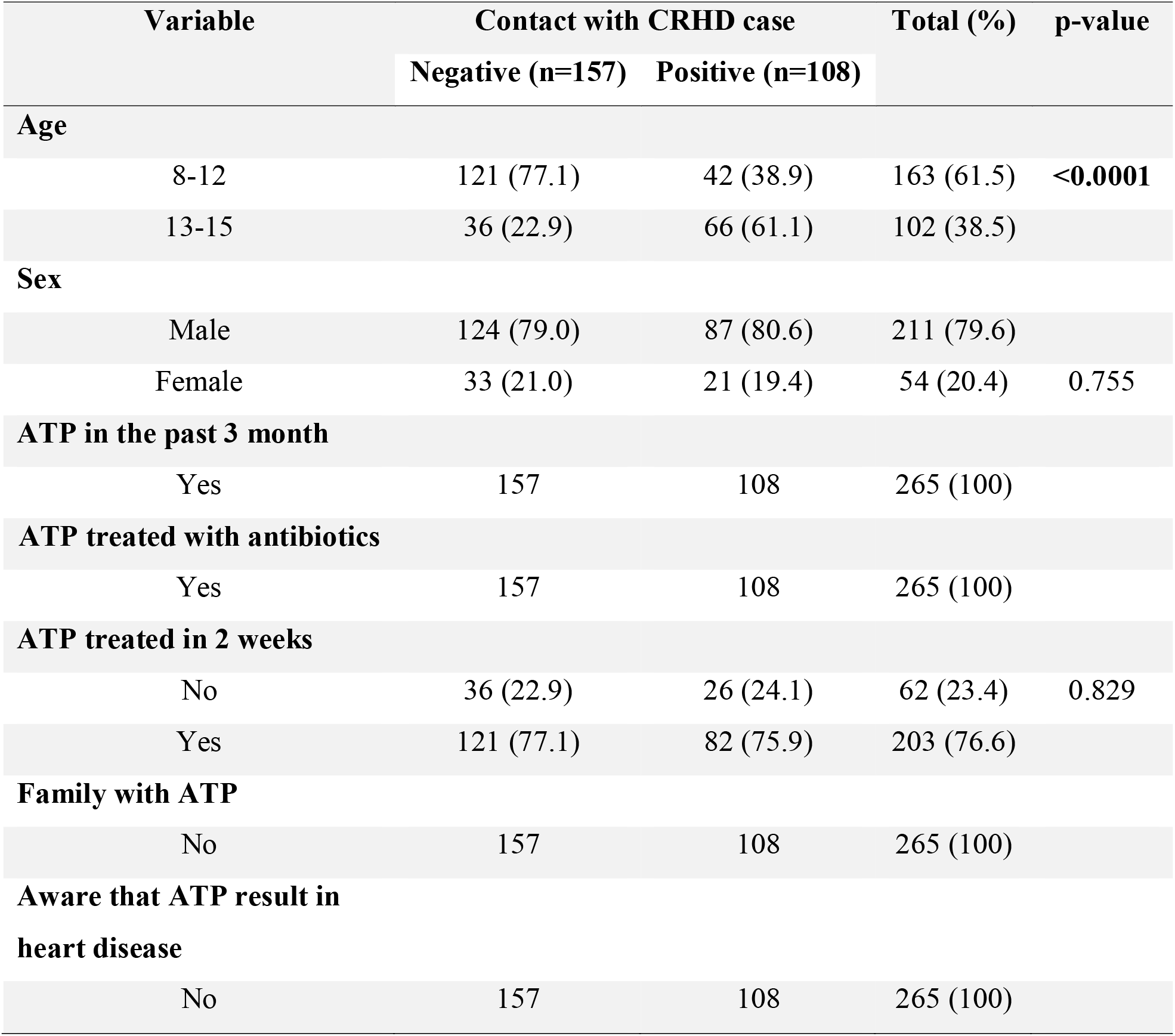
Baseline characteristics of the participants and comparison between groups (n=265)

### Total and Stratified Prevalence of Asymptomatic RHD

Among the total 265 participants, 13 were diagnosed to have asymptomatic RHD, resulting in an overall prevalence 4.9% (95% CI= 2.6-7.9%). From the 13 cases, 3 were borderline and 10 were definitive cases of RHD.

The stratified prevalence of asymptomatic RHD among negative contacts was 3/157 (1.9%, 95% CI= 0 – 4.0%) of which 2 were borderline cases. All cases were having mild MR with only one of them having morphologic mitral valve change which is thickened Anterior Mitral Valve Leaflet (AMVL) measured 4mm. There was no aortic valve lesion identified.

The prevalence among positive contacts was 10/108 (9.3%, 95% CI= 4.6 – 14.8%) of which 9 were definitive cases of RHD. Mild mitral regurgitation was seen in 8 students and mild to moderate in 2 students. There were no cases of pathologic aortic regurgitation. There was one case with pathologic mitral valve morphology with thickened AMVL but normal paravalvular structures.

Further statistical comparison to see the difference in the prevalence between the two groups shows that the difference in prevalence is also significant with a p-value of 0.003. (Table 2)

**Table 2:**
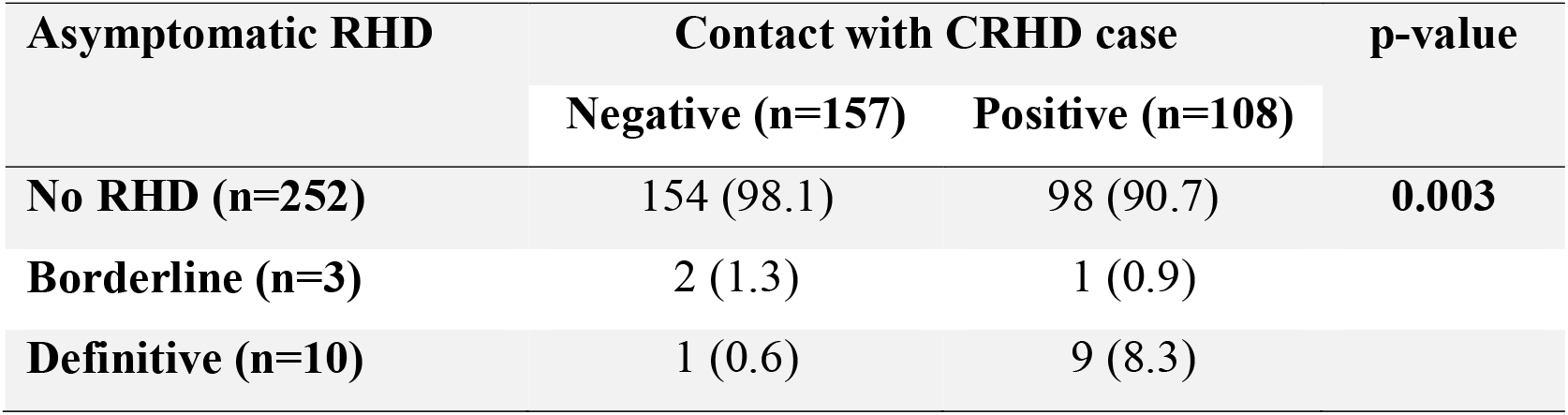
Comparison of diagnosis of asymptomatic RHD between negative and positive contacts (n=265)

**Table 3:**
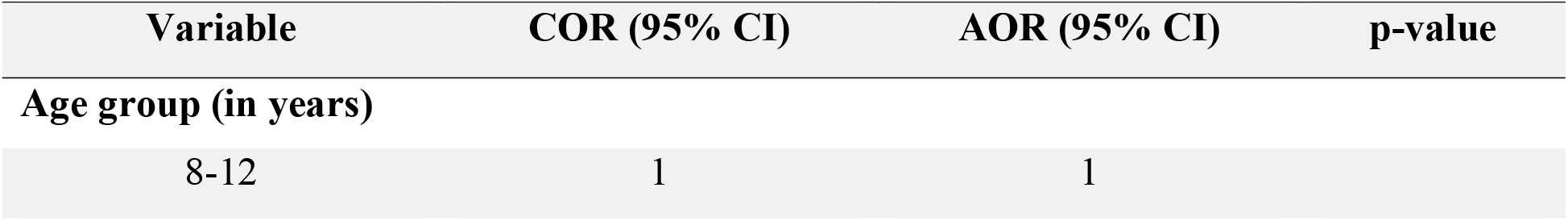
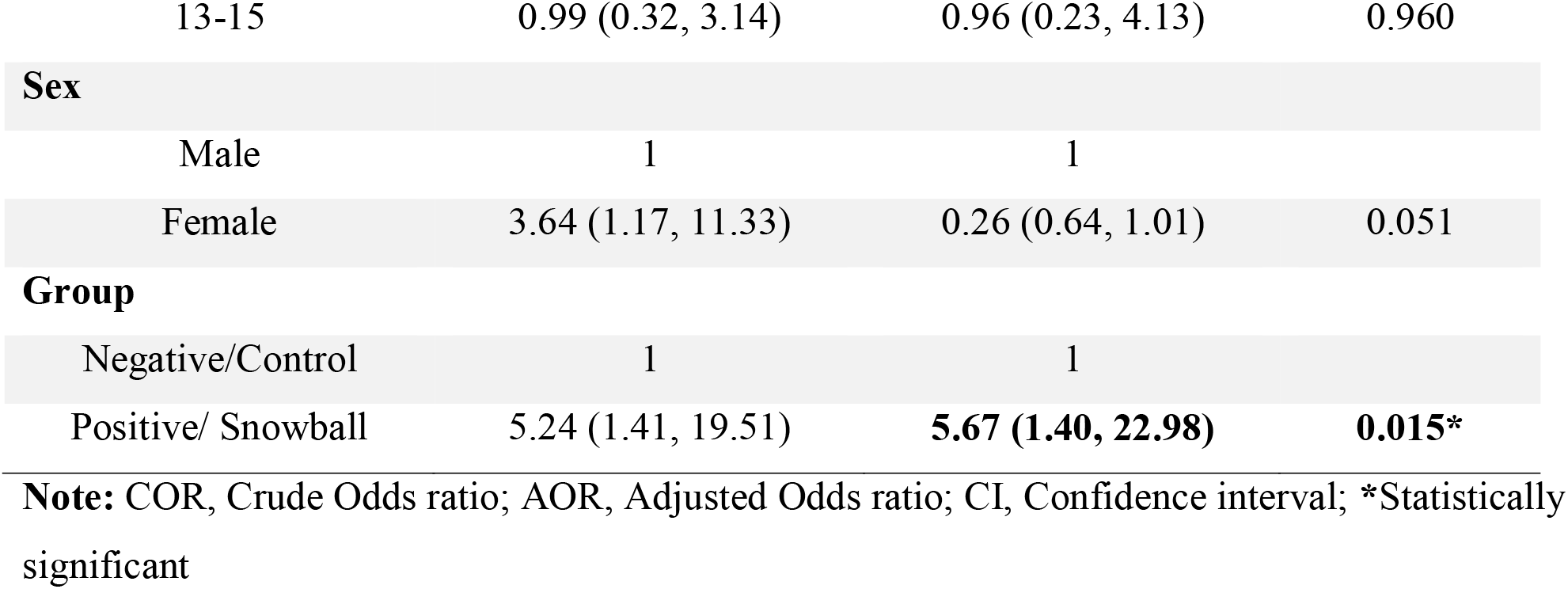
Factors associated with development of asymptomatic RHD among children (n=265)

| Variable | COR (95% CI) | AOR (95% CI) | p-value |
| --- | --- | --- | --- |
| **Age group (in years)** |  |  |  |
| 8-12 | 1 | 1 |  |
| 13-15 | 0.99 (0.32, 3.14) | 0.96 (0.23, 4.13) | 0.960 |
| **Sex** |  |  |  |
| Male | 1 | 1 |  |
| Female | 3.64 (1.17, 11.33) | 0.26 (0.64, 1.01) | 0.051 |
| **Group** |  |  |  |
| Negative/Control | 1 | 1 |  |
| Positive/ Snowball | 5.24 (1.41, 19.51) | **5.67 (1.40, 22.98)** | **0.015\*** |

**Note:** COR, Crude Odds ratio; AOR, Adjusted Odds ratio; CI, Confidence interval; \*Statistically significant

### Factors associated with development of asymptomatic RHD

A multivariable binary logistic regression model was fit with age, sex and contact history and it was found a significant association between contact history and the development of asymptomatic RHD. Accordingly, after adjusting for age and sex, the odds of developing asymptomatic RHD among participants with positive contact history was 5.67 times than participants with negative contact history (AOR= 5.67, 95% CI= 1.40, 22.98, p-value=0.015).(Table 3)

## Discussion

In this study, there were fairly comparable two groups of children of negative and positive contacts with known cases of RHD from follow up clinic. The overall prevalence asymptomatic RHD was high. We also found that the prevalence of asymptomatic RHD was significantly higher among children with a positive contact history. Furthermore, the majority of children with a positive contact history were diagnosed with definitive RHD. Regression analysis showed that having a positive contact history is associated with 5.67 times increased odds of developing asymptomatic RHD as compared with those with negative contact history.

The prevalence of asymptomatic RHD among school children showed 1.9% in East Africa (16) with a range of 0.32-4.09% which is comparable to our finding except in the contact positive groups. All the studies mentioned in the meta-analysis followed simple random school screening irrespective of contact history. We used the same group and compared it with positive contact groups which had shown significantly higher prevalence.

The use of snowball approach for RHD screening is very limited. World Health Organization and WHF recommend comprehensive control program for RHD primarily through registration of cases (15). Case detection was recommended through the use of echocardiography by standard criteria. There was scarcity of data on the strategies of case detection, either through school screening program or community-based awareness sessions. Snowball approach came to practice to increase the yield of case detection so that it can save resources (15,17).

The use of definitive and borderline RHD was brought in practice for better screening and actively detecting asymptomatic RHD (6, 15). WHF criteria was set to standardize the diagnostic approaches with improved reliability and reproducibility (15). Accordingly, definitive asymptomatic RHD means children with echocardiographic findings of pathologic mitral regurgitation with at least two morphologic features of RHD of the mitral valve or aortic valve or signs of mitral stenosis. Whereas borderline RHD was defined as having morphologic features of RHD of the mitral valve with no pathologic feature.

Our findings can be explained by the pathophysiology of development of acute rheumatic fever and RHD. Acute rheumatic fever develops from sore throat secondary to untreated GABHS infection. This bacterial sore throat is highly contagious through aerosol droplets which is high likely in the school setting of developing nations. Irrational use of antibiotic plays significant role for poor primary prevention and further resistance to antibiotics in the developing countries like Ethiopia (18).

When interpreting these findings, it is important to consider both its strengths and limitations. The limitation of the study was that additional variables that could determine development of RHD were not collected in depth. And the sample size was relatively small, which led to the lower power of the study. To improve the yield, we used standardized and structured method of case detection in both groups The two groups were fairly comparable to reach to a conclusion with rigorous statistical analysis and quality of data.

## Conclusion

The detection of asymptomatic RHD using a snowball approach to close contacts and classmates of known cases of RHD is superior to the routine school based random screening. Therefore, a selective national screening system can be designed for an effective and efficient method of identifying cases. Further detailed study with increased sample size is recommended.

## Data Availability

All relevant data are available upon reasonable request.

## Declaration

### Ethical consideration

The study was conducted after obtaining ethical clearance from St. Peter specialized hospital Institutional Review Board. Permission from the local educational office in Addis Ababa and the schools was obtained. Oral consent was obtained from the students’ parents/legal guardians and the students themselves. The study had no risk/negative consequence on those who participated in the study. Positive cases identified were linked to the hospital for treatment and further regular follow up. Study identification numbers were used for the data collection and personal identifiers were not used in the research report. Access to the collected information was limited to the principal investigator and confidentiality was maintained throughout the project.

### Availability of data and materials

All relevant data are available upon reasonable request.

### Author’s Contribution

DM conceived and designed the study, and performed echocardiography with assistance from TM. TWL contributed to the design of the study. DM and TWL performed statistical analysis, and drafted the initial manuscript, all other team members critically reviewed and contributed for approval of the manuscript. all authors revised the manuscript and approved the final version.

## Acknowledgement

The authors would like to thank St. Peter specialized hospital, woreda 8 education bureau of Gulele sub-city, the schools and data collectors for their support and facilitation of the research work.

